# Art therapy as treatment modality for persons living with dementia- a protocol for a scoping review of systematic reviews

**DOI:** 10.1101/2022.04.14.22273883

**Authors:** Elena Guseva, Machelle Wilchesky

**Author notes:** **Corresponding author:** Machelle Wilchesky. **Funding** This scoping review was funded by the Maimonides Medical Research Foundation and by Team 11 of the Canadian Consortium on Neurodegeneration in Aging (CCNA). **Author contribution** Each named author has substantially contributed to conceptualization of the underlying research and drafting this manuscript. E.G.: Conceptualization, Writing-Original draft preparation. M.W.: Writing-Review and Editing, Supervision.

## Abstract

**Objective:** To establish a robust understanding of the state of the evidence on the effectiveness and/or efficacy of art therapy (AT) as a non-pharmacological treatment (NPT) modality for persons living with dementia (PLWD).

**Background:** Over the past decade, AT has received increased attention from health care professionals and researchers as having a potential role to play within treatment plans for PLWD.(1-4)

**Inclusion criteria:** This scoping review will include systematic reviews from health-related disciplines conducted within the last 20 years that report the effectiveness and/or efficacy of AT as an NPT modality for PLWD and Mild Cognitive Impairment (MCI) as their main objective. Study outcomes must include cognition, quality of life, emotional and psychological well-being, and/or neuropsychiatric symptoms (NPS).

**Methods:** A scoping review of systematic reviews was selected to outline different types of evidence and to identify gaps in the literature. The proposed review will be guided by the methodological framework proposed by the Joanna Briggs Institute. We will therefore specify the research question, identify relevant studies, select eligible studies, extract, collate, and summarize our results. We will not conduct a quality appraisal of the included studies as this review aims to explore the general scope of research conducted that assesses AT effectiveness and efficacy in PLWD.

## Introduction

The World Health Organization recognizes dementia as being a global public health crisis of the 21^st^ century(5, 6) affecting 47.5 million people worldwide. More than 500,000 Canadians are living with Alzheimer’s disease or other forms of dementia today and this number is expected to reach 912,000 by the year 2030.(7) Dementia is a syndrome of cognitive impairment that affects memory, cognitive abilities and behaviour, and significantly interferes with a person’s ability to perform daily activities.(8) Neuropsychiatric symptoms (NPS) of dementia, that often prevalent and associated with severity, are among the most distressing sequelae of the disease.(9) Pharmacological treatments for symptoms management have been shown to be associated with adverse and potentially serious side effects,(10) and are therefore not recommended as first-line therapy.(11) The last few decades have therefore seen a proliferation in expanding the range of non-pharmacological treatments (NPTs) that aim to enhance quality of life, emotional and psychological wellbeing and reduce NPS in PLWD.

Art therapy (AT) is a psychosocial intervention that combines psychotherapy and the creative process, facilitating self-exploration and understanding.(12) Previously AT research has demonstrated its effectiveness in treatment of affective disorders and other mental health conditions in various populations and considered to be a safe and reliable non-pharmacological treatment option.(13) Art therapists advocate that art can be beneficial to individuals affected by dementia as it facilitates self-expression through sensory stimulation, creativity, and social interaction.(2, 4) Artmaking and creativity have capacity to increase ability to identify and express emotions in PLWD.(14) Usefulness of AT as an effective rehabilitative strategy for people suffering from dementia remains highly debated.(15, 16) Research suggests, however, that PLWD can produce and appreciate visual art and that aesthetic preferences can remain stable, despite cognitive decline.(16, 17) A preliminary search of MEDLINE, the Cochrane Database of Systematic Reviews and JBI Evidence Synthesis was conducted and no current or underway systematic reviews or scoping reviews on the topic were identified.

## Review question

**Our research question is:** What evidence from systematic reviews (SRs) exists on the effectiveness and/or efficacy of AT for PLWD?

## Inclusion criteria

### Participants

The current scoping review will include systematic reviews pertaining to PLWD. Studies that investigate participants of all ages, sex, levels of dementia severity, levels of comorbidity, medication use and care settings. Systematic reviews on other neurodegenerative diseases will be excluded.

### Concept

The concept examined by this scoping review is AT as NPT for PLWD and its effectiveness and efficacy on measured outcomes in this population. There are multiple and often conflicting definitions of AT that has contributed to some confusion regarding the profession itself. AT originated in United Kingdom in the late 40s as a therapeutic application of image making.(18) The British Art Therapy Association defines AT is “a form of psychotherapy that uses art media as its primary mode of expression and communication. Within this context, art is not used as diagnostic tool but as a medium to address emotional issues which may be confusing and distressing”.(19) According to the Canadian Art Therapy Association, AT “combines the creative process and psychotherapy, facilitating self-exploration and understanding. Using imagery, colour and shape as part of this creative therapeutic process, thoughts and feelings can be expressed that would otherwise be difficult to articulate”.(12) The American Art Therapy Association states that AT “is an integrative mental health and human services profession that enriches the lives of individuals, families, and communities through active art-making, creative process, applied psychological theory, and human experience within a psychotherapeutic relationship.”(20) AT may be administered either individually or via group sessions by professional licensed art therapists.

AT as a professionally led modality can be differentiated from participatory arts programmes such as arts and crafts classes, attending museums or galleries, painting and sculpting and many other activities that employ visual arts.(21) The key difference is that AT must be delivered by trained professional therapists and have specific psychotherapeutic aims, whereas arts in healthcare settings or in everyday life are commonly delivered by artists or other individuals who do not have specific therapy training.(21) Although there remains debate pertaining to the similarities and differences of both approaches, the involvement of a trained therapist remains the defining distinction separating AT from other art activities.(21)

We anticipate that some included systematic reviews will not meet this AT definition and will therefore include studies that do not qualify as AT studies per se, despite reporting otherwise. Given that this scoping review is a precursor for a larger planned AT systematic review, this scoping review will overlook strict definitions of AT and will include systematic reviews that investigate any visual arts-based approaches and interventions.

### Context

The current scoping review will consider systematic reviews involving PLWD as participants who reside either in the community or in the residential care facility setting. Studies from any geographic location will be eligible for inclusion.

### Types of Sources

This scoping review will consider any systematic review reporting on the effectiveness and/or efficacy of AT as an NPT modality for PLWD and persons with MCI as their main objective published after the year 2002 that meet the inclusion criteria, regardless of whether results are provided narratively or quantitatively (or both). All other types of studies will be excluded.

## Methods

The proposed scoping review will be conducted in accordance with the JBI methodology for scoping reviews.(22)

### Search strategy

The search strategy will aim to identify both published and unpublished English and French language systematic reviews. An initial limited search of MEDLINE (PubMed), CINAHL, PsycINFO, and Google Scholar will be undertaken to identify articles on this topic, followed by analysis of the text words contained in the titles and abstracts, and of the index terms used to describe these articles. This will inform the development of a search strategy including identified keywords and index terms that will be tailored for each information source. A full search strategy is provided in Appendix I. The reference lists of all included studies will be screened for additional studies.

The databases to be searched will include:

- CINAHL
- MEDLINE (PubMed)
- PsycINFO
- Embase
- Cochrane Database of Systematic Reviews
- *JBI Database of Systematic Reviews and Implementation Reports*

The search for unpublished studies will include:

- ProQuest Dissertation and Theses
- Google Scholar/Google

### Study/Source of Evidence selection

Following these searches, all identified citations will be collated and uploaded into the Covidence web-based software platform that streamlines the production of research reviews where duplicates will be removed.(23) All citation details will be imported into Covidence. Following screening criteria pilot testing, titles and abstracts of each article will then be screened by two independent reviewers for assessment against the inclusion criteria for the review. Full text of all potentially relevant sources will then be retrieved.

The full text of selected citations will be assessed in detail against the inclusion criteria by two or more independent reviewers. Reasons for exclusion of sources of evidence of full text that do not meet the inclusion criteria will be recorded and reported in the scoping review. Any disagreements that arise between reviewers at each stage of the selection process will be resolved through discussion, and unresolved conflicts will be adjudicated by the senior author (MW).

Results of our review will be reported according to the Preferred Reporting Items for Systematic Reviews and Meta-analyses extension for scoping reviews (PRISMA-ScR).(24) The reference lists of all included studies will be screened for additional relevant studies through backward and forward snowballing.

### Data Extraction

Data from studies included in this scoping review will be extracted by two independent reviewers using the draft data extraction tool that will capture specific details about the populations, concept, context, study methods, and results significant to our objectives. Any disagreements that arise between reviewers will be resolved through discussion, and unresolved conflicts will be adjudicated by the senior author (MW). Where required, authors of papers will be contacted to request missing or additional data.

The draft data extraction tool will be pilot tested and modified and revised as necessary during the process of data extraction. In line with the Joanna Briggs Institute methodology,(22) the review team can extract any additional relevant data from included studies. Any modifications to the original protocol will be detailed in the published scoping review report.

### Data Analysis and Presentation

The results of the search will be reported in full in the final scoping review report and presented in a Preferred Reporting Items for Systematic Reviews and Meta-Analyses extension for Scoping Reviews (PRISMA-ScR) flow diagram.(25) Extracted data will be presented in tabular form with a narrative summary related to the key findings that convey the objectives of the present scoping review. Due to the heterogeneity of the studies anticipated for this review, findings of the included studies will be described narratively. Data analysis and presentation may be further refined during the review process as the reviewers become more aware of the content of all of their included studies.

## Data Availability

All data produced in the present work are contained in the manuscript

## Acknowledgements

This scoping review is to contribute towards a PhD degree for E.G.

## Appendices

## Appendix I: Search strategy for Ovid MEDLINE(R) ALL <1946 to March 25, 2022>

| Search | Query | Records retrieved |
| --- | --- | --- |
| #1 | *Dementia.mp. or Dementia/* | 145,261 |
| #2 | *Alzheimer's disease.mp. or Alzheimer Disease/* | 167,307 |
| #3 | *art therapy.mp. or Art Therapy/* | 2,254 |
| #4 | *Art Therapy/ or Creative arts therapies.mp. or Creativity/* | 9,049 |
| #5 | *systematic review.mp. or "Systematic Review"/* | 250,966 |
| #6 | *1 or 2* | 262,803 |
| #7 | *3 or 4* | 9,618 |
| #8 | *6 and 7* | 286 |
| #9 | *5 and 8* | 13 |
| Limited to the studies conducted after 2002 and published in English and French languages |  |  |

## Appendix II: Data extraction instrument

|  |  |
| --- | --- |
| 1 | Study identification |
| 2 | Number of databases searched |
| 3 | Search dates |
| 4 | Search terms used |
| 5 | Number and type of included studies |
| 6 | Was AT the primary intervention? |
| 7 | Other interventions |
| 8 | Definition of AT |
| 9 | Inclusion criteria |
| 10 | Exclusion criteria |
| 11 | Main research objective |
| 12 | Dementia type/MCI |
| 13 | Severity of dementia |
| 14 | Outcomes investigated |
| 15 | Outcomes measurements |
| 16 | Reported results |

